# The UK Coronavirus Job Retention Scheme and changes in diet, physical activity and sleep during the COVID-19 pandemic: Evidence from eight longitudinal studies

**DOI:** 10.1101/2021.06.08.21258531

**Authors:** Bożena Wielgoszewska, Jane Maddock, Michael J. Green, Giorgio Di Gessa, Sam Parsons, Gareth J Griffith, Jazz Croft, Anna J. Stevenson, Charlotte Booth, Richard J. Silverwood, David Bann, Praveetha Patalay, Alun D. Hughes, Nish Chaturvedi, Laura D Howe, Emla Fitzsimons, Srinivasa Vittal Katikireddi, George B. Ploubidis

## Abstract

**Background:** In March 2020 the UK implemented the Coronavirus Job Retention Scheme (furlough) to minimize job losses. Our aim was to investigate associations between furlough and diet, physical activity, and sleep during the early stages of the COVID-19 pandemic.

**Methods:** We analysed data from 25,092 participants aged 16 to 66 years from eight UK longitudinal studies. Changes in employment (including being furloughed) were defined by comparing employment status pre- and during the first lockdown. Health behaviours included fruit and vegetable consumption, physical activity, and sleeping patterns. Study-specific estimates obtained using modified Poisson regression, adjusting for socio-demographic characteristics and pre-pandemic health and health behaviours, were statistically pooled using random effects meta-analysis. Associations were also stratified by sex, age, and education.

**Results:** Across studies, between 8 and 25% of participants were furloughed. Compared to those who remained working, furloughed workers were slightly less likely to be physically inactive (RR:0.85, [0.75-0.97], I^2=^59%) and did not differ in diet and sleep behaviours, although findings for sleep were heterogenous (I^2=^85%). In stratified analyses, furlough was associated with low fruit and vegetable consumption among males (RR=1.11; 95%CI: 1.01-1.22; I^2^: 0%) but not females (RR=0.84; 95%CI: 0.68-1.04; I^2^: 65%). Considering change in these health behaviours, furloughed workers were more likely than those who remained working to report increased fruit and vegetable consumption, exercise, and hours of sleep.

**Conclusions:** Those furloughed exhibited broadly similar levels of health behaviours to those who remained in employment during the initial stages of the pandemic. There was little evidence to suggest that such social protection policies if used in the post-pandemic recovery period and during future economic crises would have adverse impacts on population health behaviours.

## Introduction

COVID-19 disease, social distancing measures and a series of lockdowns have affected the economy and employment rates in the United Kingdom (UK) and worldwide (1,2). The current pandemic has also resulted in health care disruptions and closures of some sectors of the economy including exercise facilities. This unique situation makes it difficult to predict the short and long-term effects of pandemic-related unemployment on population health and health related behaviours.

Social protection policies introduced during the pandemic may modify the health consequences of the COVID-19-related economic downturn. The UK Government launched their Coronavirus Job Retention Scheme (CJRS) in March 2020. The CJRS, widely referred to as ‘furlough’, provides employees unable to work due to the pandemic with 80% of pay (capped at £2,500 per month) (3). By March 2021, 11.4 million employees (approximately 34% of those over 16 years in employment) had been furloughed through the CJRS and the number of people claiming unemployment-related benefits had increased by 1.4 million from March 2020 (4). These economic changes have not affected all groups equally. Younger workers, low earners and women, were more likely to work in disrupted sectors, and therefore become unemployed or be furloughed (4,5).

The relationship between government interventions, particularly those focused on mitigating the impact of lockdown and economic downturns via subsidised employment, and health is poorly understood. The CJRS may impact health through its influence on health behaviours. We aimed to investigate associations between changes in employment status (with a focus on the UK’s furlough scheme) during the early stages of the pandemic and health behaviours namely diet, physical activity and sleep by conducting coordinated analyses of data from more than 25,000 participants in eight longitudinal studies. We hypothesised that associations differ by participant characteristics, therefore we also examined associations stratified by sex, education, and age.

## Methods

### Participants

The UK National Core Studies Longitudinal Health and Wellbeing initiative is drawing together data from multiple UK population-based longitudinal studies using coordinated analysis to answer priority pandemic-related questions. By conducting similar analyses within each study and pooling results in a meta-analysis, we can provide robust evidence to understand how the pandemic has impacted population health and support efforts to mitigate its effects going forward. Data here were from eight long running UK population-based longitudinal studies which conducted surveys during the pandemic. Details of the design, sample frames, current age range, timing of the most recent pre-pandemic and COVID surveys, response rates, and sample size are in Supplementary Table S1.

Five studies were age homogenous birth cohorts (where all individuals within each study were similar age): the Millennium Cohort Study (MCS); the index children from the Avon Longitudinal Study of Parents and Children (ALSPAC-G1); Next Steps (NS, formerly the Longitudinal Study of Young People in England); the 1970 British Cohort Study (BCS70); and the 1958 National Child Development Study (NCDS). Three age heterogeneous studies (each covering a range of age groups) were included: Understanding Society (USOC); the English Longitudinal Study of Ageing (ELSA); and Generation Scotland: the Scottish Family Health Study (GS). Finally, the parents of the ALSPAC-G1 cohort were treated as a fourth age heterogeneous study population (ALSPAC-G0).

Analytical samples were restricted to working age participants, defined as those aged 16 to 66 (the current state pension age in the UK(6)), who had recorded at least one health behaviour outcome in a COVID-19 survey and had valid data on all covariates. Most studies were weighted to restore representativeness to their target populations, accounting for sampling design where appropriate and differential non-response to pre-pandemic and COVID surveys (7). Weights were not available for GS. Details of the weighting applied within each study are in Supplementary Table S1.

### Measures

Below we describe all variables in the analysis. Full details of the questions and coding used within each cohort are in Supplementary File 2.

### Exposure: Employment status change

Employment status change (or stability) was coded in six categories based on the status both prior to the pandemic and at their first COVID-19 survey: stable employed (reference group); furloughed (i.e. from work to furlough); no longer employed (i.e. from employed to non-employed); became employed (i.e. from non-employed to employed); stable unemployed (i.e. unemployed at both points); and stable non-employed (i.e. not available for employment at either point, including in education, early retirement, caring responsibilities, sick or disabled).

### Outcomes: Health behaviours

We examined diet, physical activity, and sleep. Participants self-reported fruit and vegetable consumption (≤2 portions per day vs more portions (8)), time spent exercising (<3 days a week for 30 minutes or more vs more frequent exercise within recommended levels (9)), and hours of sleep (outside the typical range of 6-9 hours vs within that range (10)) both during and pre-pandemic. However, this information, used for our main analyses, was only available in some studies (MCS, NS, BCS, NCDS, USOC), whereas others (ALSPAC, GS, ELSA) only had information on change since the start of the pandemic (see Supplementary File 2). Based on these levels *or* on the information on changes in health behaviours since the start of the pandemic, we additionally created dichotomous outcomes indicating change from before to during the pandemic (in comparison to no change or change in the other direction): more portions of fruit/vegetables; fewer portions of fruit/vegetables; more time spent exercising; less time spent exercising; more hours of sleep; fewer hours of sleep; a shift from outside to within the typical sleep range of 6-9 hours; and a shift from within to outside the typical sleep range of 6-9 hours. All information on behaviours during the pandemic was from surveys conducted between April and July 2020 (inclusive).

### Confounders and Moderators

Potential confounders included: sex; ethnicity (non-white ethnic minority vs white -including white ethnic minorities); age; education (degree vs no degree); UK nation (i.e. England, Wales, Scotland, Northern Ireland or other); household composition (based on presence of a spouse/partner and presence of children); pre-pandemic psychological distress; pre-pandemic self-rated health (excellent-good vs fair-poor); and pre-pandemic health behaviour measures, where available.

We examined modification of the associations by sex, education, and age in three categories: 16-29; 30-49; and 50 years or more (with age-homogeneous cohorts included in the relevant band).

### Analysis

Within each study, each outcome was regressed on employment status change, using a modified Poisson model with robust standard errors that returns risk ratios for ease of interpretation and to avoid issues related to non-collapsibility of odds ratios (11,12). After estimating unadjusted associations, confounder adjustment was performed in two steps. First, a “basic” adjustment including socio-demographic characteristics: age (only in age-heterogeneous studies), sex, ethnicity (except the BCS and NCDS cohorts which were nearly entirely white), education, UK nation (except ALSPAC, GS and ELSA which only had participants from a single country), and household composition. Second, a “full” adjustment additionally including pre-pandemic measures of: psychological distress, self-rated health, and health behaviours. Moderation by sex, age, and education was assessed with stratified regressions using “full” adjustment.

Both stages of adjustment are relevant because our exposure, employment change, incorporates pre-pandemic employment status, which may have influenced other pre-pandemic characteristics such as mental health, self-rated health, and health behaviours (see supplementary Figure S8). By not controlling for these pre-pandemic characteristics, the basic adjusted risk ratios may represent both newly acquired behaviour and/or continuation of established (pre-pandemic) behaviour. In contrast, the full adjustment risk ratios block effects via these pre-pandemic characteristics and can therefore be interpreted as representing the differential change in health behaviour between exposure groups which is independent of these pre-pandemic characteristics. For the outcomes that directly capture changes in health behaviour, the full adjustment did not include pre-pandemic levels of the behaviour in question, as pre-pandemic levels of that behaviour are incorporated within the change outcome. This means that even full adjustment risk ratios estimated for these outcomes may partially reflect associations with pre-pandemic behaviour.

The overall and stratified results from each study were pooled using a random effects meta-analysis with restricted maximum likelihood in Stata 16. For stratified results, a test of group differences was performed using the subgroup meta-analysis command. Some studies could not contribute estimates for every comparison due to differences in the ages sampled, measures used, and sparsity of data. For a small number of exposure-outcome comparisons, reliable estimates could not be computed because the outcome prevalence was low (≤2). While such selective exclusion could potentially lead to bias, the low numbers of events mean that the corresponding within-study estimates would be so imprecise that their exclusion is unlikely to lead to considerable bias (see Supplementary File 3 for more details and sensitivity analyses showing that results were robust to different low cell count exclusion thresholds). We report heterogeneity using the I^2^ statistic: 0% indicates estimates were similar across studies, while values closer to 100% represent greater heterogeneity. While we could have undertaken multivariate meta-analysis of all exposure categories simultaneously, for ease of interpretation we instead conducted a series of univariate meta-analyses, bearing in mind the consistency of results from these approaches generally observed elsewhere (13,14). We performed a multivariate meta-analysis with one outcome in a subset of the studies as a sensitivity analysis, and differences from the individual univariate meta-analyses were negligible (results not shown).

## Results

Analyses included 25,092 individuals from eight studies (see Supplementary Table S3 for demographic characteristics). Figure 1 shows employment status change during the first lockdown of the pandemic. Around six in 10 participants in NS, BCS, GS, USOC, and ALSPAC were employed prior to and during the initial stages of the pandemic, with the younger (MCS) and older studies (ELSA and NCDS) showing lower levels of stable employment. Prevalence of furlough ranged between 8% (GS) and 25% (NS). Across most studies approximately 3% of participants were no longer employed during the pandemic (8% in ALSPAC G0). Stable unemployment ranged in prevalence between 1% (GS) and 9% (ALSPAC G0). Supplementary Table S4 shows how economic activity was patterned by education, sex, and age-groups, with furlough generally more common among younger, female and less educated participants and stable employment especially common among male, higher educated and middle-aged participants. There were no clear patterns across studies with regard to who was no longer employed during the pandemic.

**Figure 1:**
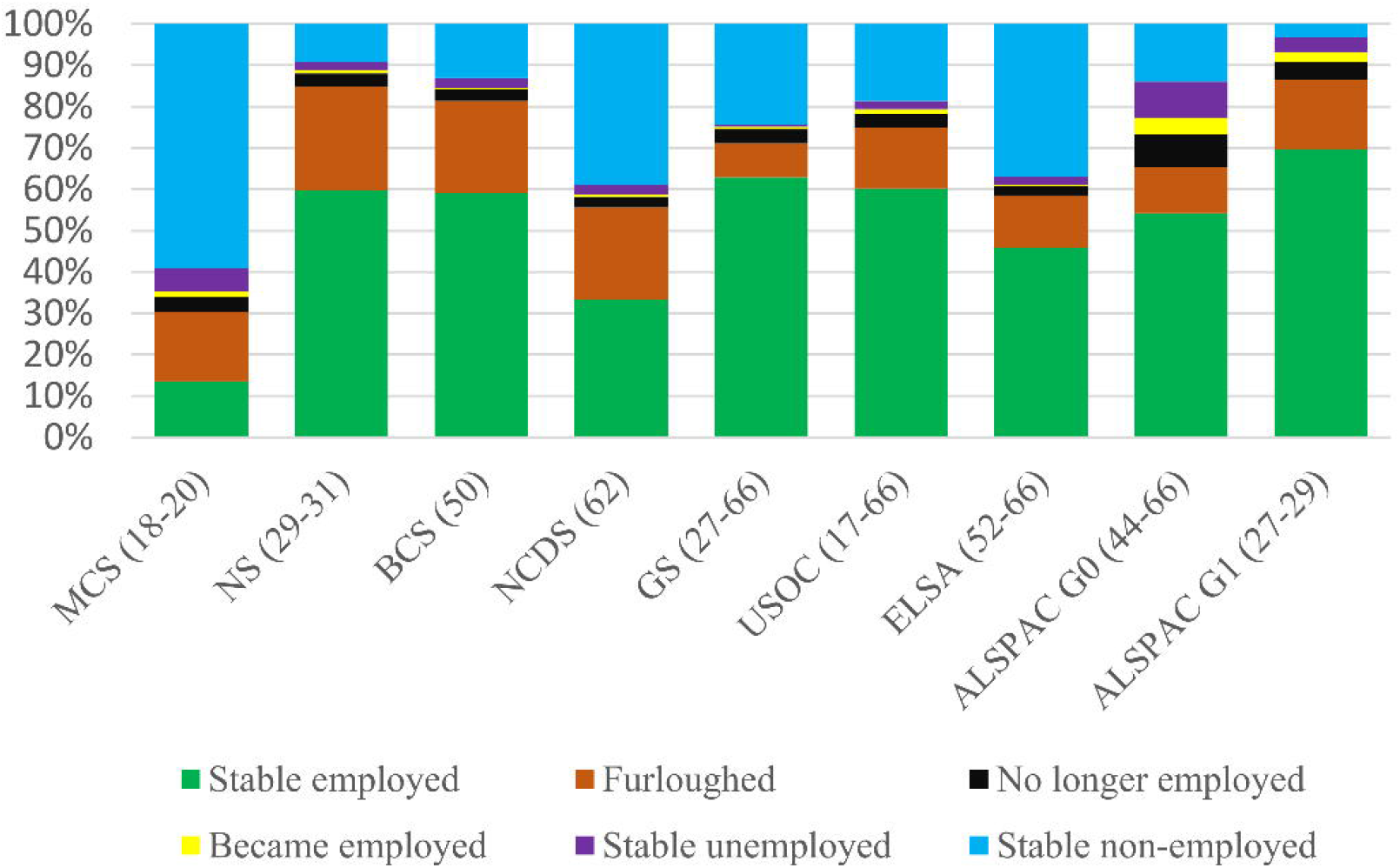
Percent distribution of change in employment status during the pandemic by study. Supplementary Table 1 has details of each study’s sample design and weighting applied. Analysis for GS, USOC, and ELSA restricted to participants aged 66 and younger. For more information about the questions asked in each dataset to derive changes in economic activity, please see Supplementary File 2.

Table 1 shows the prevalence of health behaviours and changes in behaviour by study. Proportions reporting eating no more than 2 portions of fruit or vegetables per day and reporting three or less days a week with at least 30 minutes of exercise were similar both pre- and during the pandemic, whereas sleep outside of the typical range of 6-9 hours was more common during the pandemic in most studies (USOC was an exception). Nevertheless, changes in all three behaviours were common in both directions. In the four national birth cohorts (which used identical questions), more participants reported increasing their fruit and vegetable consumption and exercise than those who reported decreases, while reporting more hours of sleep and shifts to sleep outside the typical range, were more common than reporting fewer hours of sleep, or shifts from outside to within the typical range.

**Table 1:**
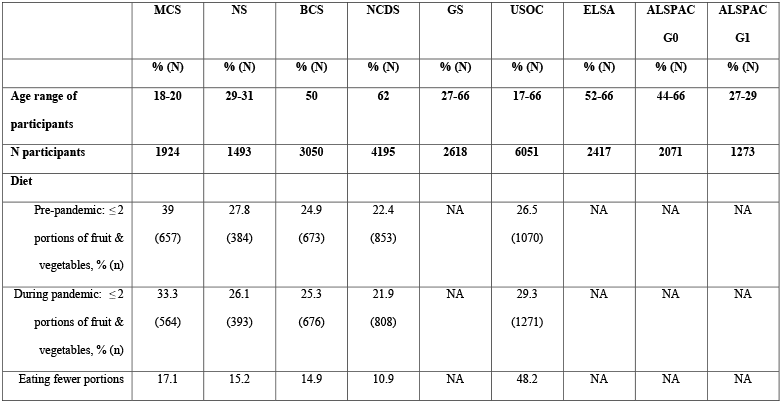
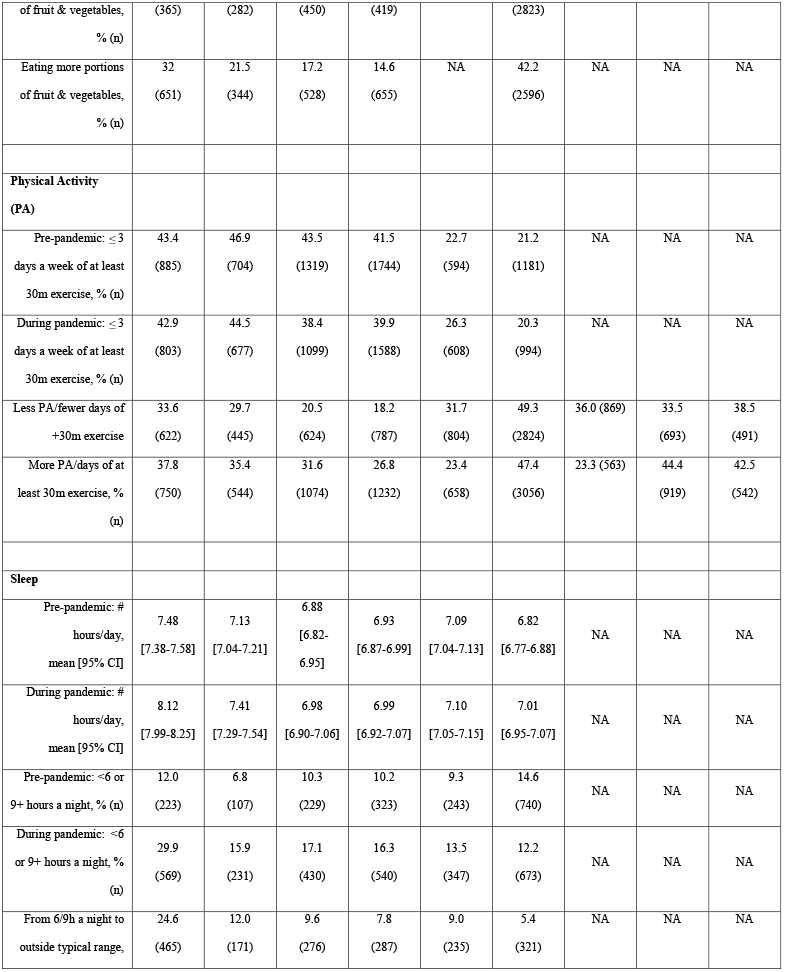
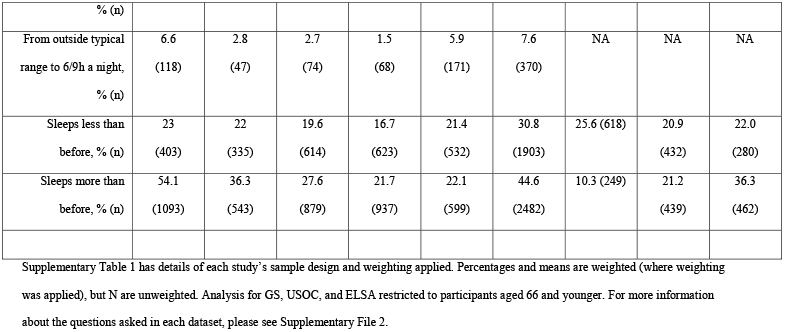
Percent (and N) distribution of health behaviours and changes during the pandemic by study.

### Pooled Analysis

Figure 2 shows meta-analysis estimates from unadjusted, basic adjusted, and fully adjusted models for levels of fruit and vegetable consumption, physical activity, and sleep during the pandemic. Given our primary interest in investigating health behaviours of those furloughed, no longer employed and stable unemployed compared to those in stable employment, we only present results for these groups (omitting those who became employed or were in stable non-employment). Figure 3 shows pooled estimates from fully adjusted models stratified by sex, education and age. Stratified estimates were largely consistent with the main results, though we highlight some differences below. Full details of the meta-analysis including overall and stratified estimates from each study are available in Supplementary File 3.

**Figure 2:**
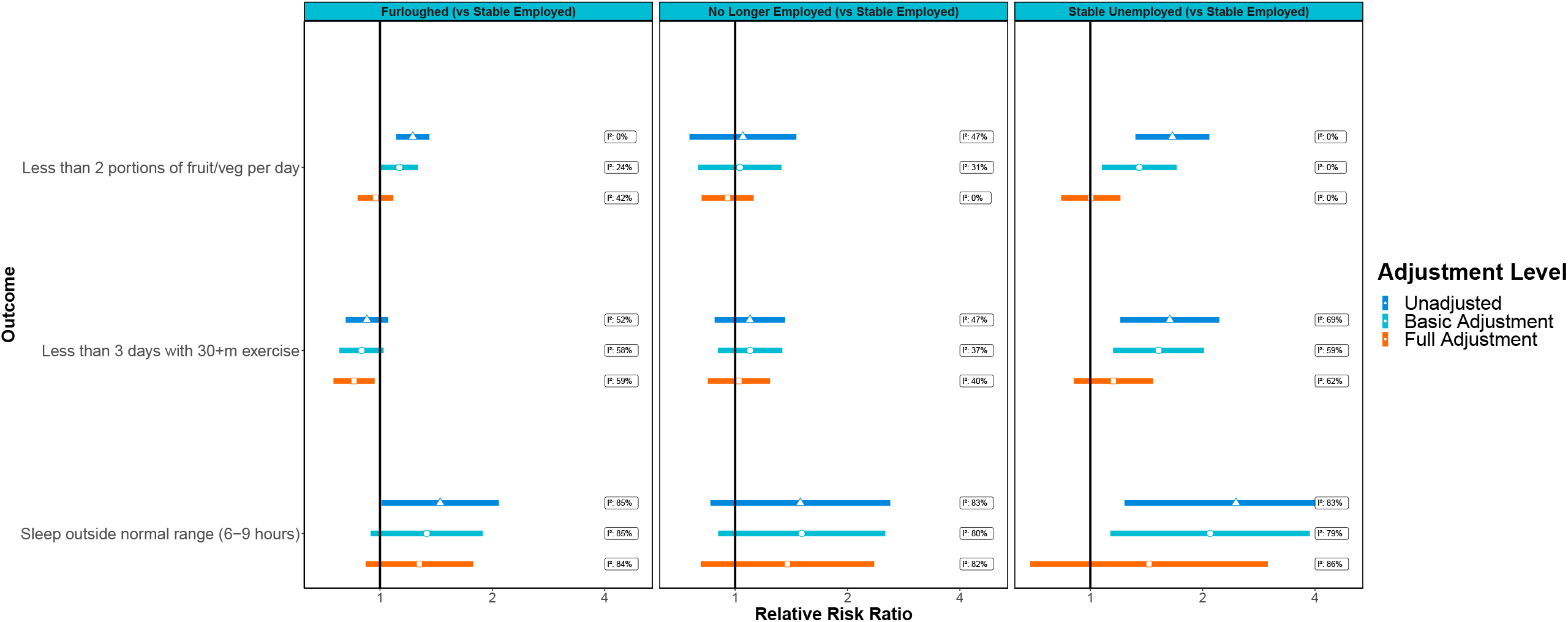
Associations between economic activity and health behaviours in pooled analyses across eight UK longitudinal studies. ‘Basic’ adjustment includes age, sex, ethnicity, education, UK nation, and household composition. ‘Full’ adjustment additionally includes pre-pandemic measures of mental health, self-rated health, diet, exercise and sleep.

**Figure 3:**
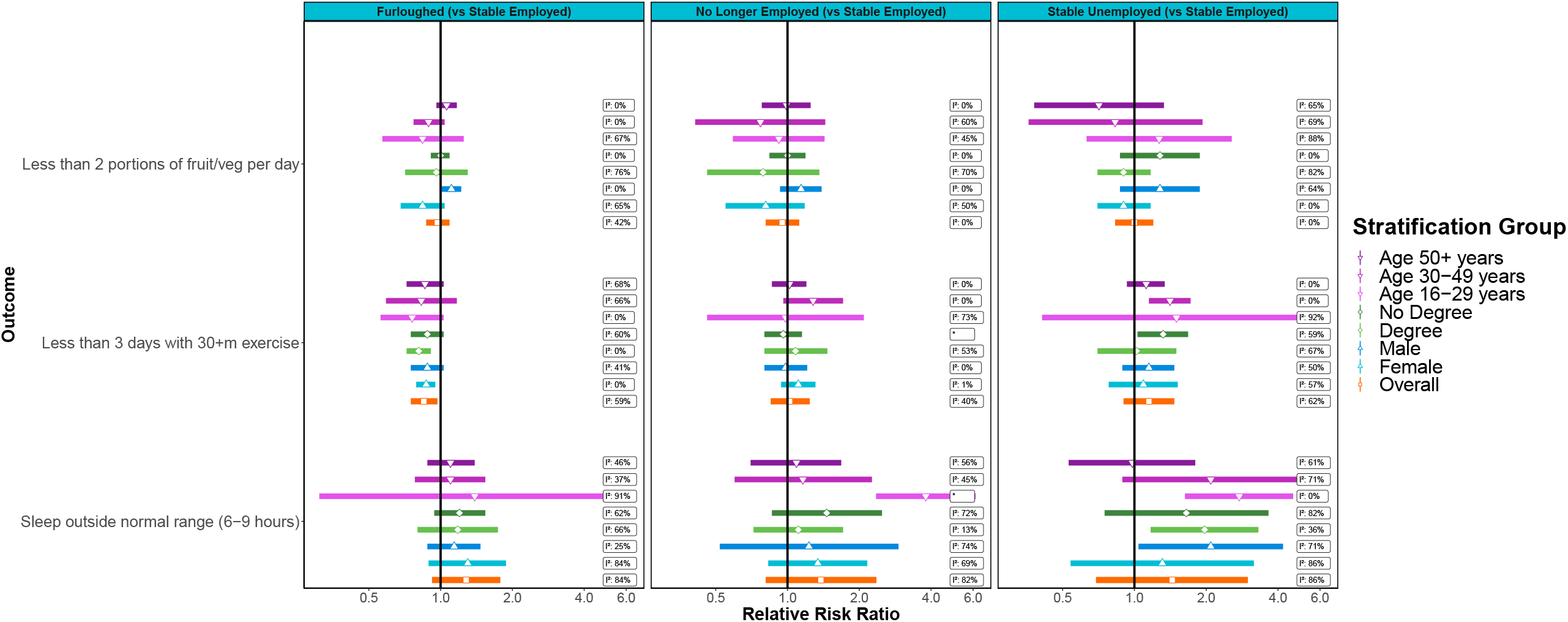
Associations between economic activity and health behaviours stratified by age, sex and educational attainment. *No I^2^ value as only one study was able to provide an estimate.

### Fruit and vegetable consumption

Unadjusted estimates suggest lower fruit and vegetable consumption during the pandemic among those furloughed or in stable unemployment compared to those who remained employed. These differences were robust to the basic adjustment, but were attenuated with full adjustment for pre-pandemic characteristics, suggesting that these associations are attributable to differences in dietary habits established prior to the pandemic. We observed moderate heterogeneity in the fully adjusted furlough model (I^2^=42%). When looking at individual studies, only associations in MCS, where participants were 18-19 years old, remained after full adjustment (RR=0.68; 95%CI: 0.49 to 0.69, 9% of the overall estimate, Supplementary File 3). There were no clear overall differences in fruit and vegetable consumption between those in stable employment and those who were no longer employed during the pandemic.

The association between furlough and fruit and vegetable consumption differed by gender (*p*=0.02). Males who were furloughed were more likely to consume less fruit and vegetables during the pandemic than males who remained employed (RR=1.11; 95%CI: 1.01-1.22; I^2^: 0%). This association was not observed among furloughed females (RR=0.84; 95%CI: 0.68-1.04), although there was heterogeneity (I^2^=65%) and furloughed females from MCS (again, the clearest outlier) were less likely to have low fruit and vegetable consumption than MCS females remaining employed (RR=0.51; 95%CI: 0.34-0.77). We did not observe differences by education or age.

### Physical Activity

Compared to stable employment, furlough was associated with lower risk of infrequent physical activity in fully adjusted models. In contrast, estimates for those no longer employed or in stable unemployment were in the opposite direction, although confidence intervals included the null. There was little evidence of subgroup differences in these associations.

### Sleep

All three groups, furloughed, no longer employed, and stable unemployment, were more likely than those in stable employment to have atypical sleep. These associations were partly attenuated in the basic adjustment and further attenuated in the full adjustment models, so were at least partially accounted for by pre-pandemic characteristics and behaviours. Estimates for sleep exhibited high heterogeneity with I^2^ values largely over 80%, perhaps partly due to age differences between the samples (see below).

The heightened risk of atypical sleep for those not in stable employment appeared to be largely concentrated at younger ages. For example, stable unemployment was associated with an RR of 2.75 (95%CI: 1.63-4.63; I^2^: 0%) in the 16-29 year age group, compared with 0.98 (95%CI: 0.53-1.80; I^2^: 61%) in the 50+ age group (p=0.04). Age patterning was similar for those no longer employed (p<0.01), but considerably less pronounced for furlough (p=0.96). Thus, in this youngest age group, even after adjusting for pre-pandemic characteristics, there was evidence that atypical sleep was associated with stable unemployment (see above for RR) or being no longer employed (RR=3.80; 95%CI: 2.35-6.15; single estimate from MCS), but there was not a clear association with furlough (RR=1.39; 95%CI: 0.31-6.16; I^2^: 91%). The two studies that had provided estimates for furlough in this age group had shown very different findings (raised risk in MCS but lower risk in USOC).

### Changes in Behaviour

Pooled estimates for the outcomes indicating change in behaviour are presented in Supplementary Tables S5, S6 and S7. These analyses indicated that furlough was associated with increases in fruit and vegetable consumption (RR=1.22; 95%CI: 1.04-1.43; I^2^: 52%), time spent exercising (RR=1.18; 95%CI: 104-1.35; I^2^: 75%) and hours of sleep (RR=1.62; 95%CI: 1.39-1.90; I^2^: 80%) relative to stable employment. Furlough was also associated with a higher likelihood of shifts both into and out of the typical 6-9 hour sleep range, which is probably due to the strong association with increased hours of sleep (which was present in all stratified analyses). These associations were robust to adjustment for other pre-pandemic characteristics, though, by the nature of change outcomes, may still partially represent pre-pandemic differences in each behaviour. Largely similar patterns were seen for: sleep among those no longer employed or in stable employment; and for physical activity among those no longer employed.

## Discussion

We find little evidence that furlough was associated with worse health behaviours. Those who were furloughed did not differ in fruit and vegetable consumption or sleep and had a lower likelihood of infrequent exercise compared to those who remained employed. Stratified analyses showed that furloughed men, but not women, had a higher likelihood of low fruit and vegetable consumption than those who remained employed. Those who remained unemployed had worse health behaviours relative to the stable employed, although these differences were largely due to pre-pandemic behaviours. Among 16-29 year olds, who were no longer employed or remained unemployed, there was a higher risk of atypical asleep.

Previous studies on subsidised employment policies have shown beneficial effects (15). Evidence from Sweden (16) shows that individuals in subsidised employment occupied an intermediate position in terms of subjective well-being; they were better-off than unemployed individuals, but worse-off than those in regular employment. Here, we have only observed minor differences between those furloughed and those in stable employment, which may be due to the nature of the CJRS scheme and/or differences in the outcomes studied. Studies conducted since the COVID-19 pandemic have shown that for some, health behaviours improved while for others they declined (17,18), and our findings offer little evidence of furlough contributing to declines in healthy behaviour.

Unemployment has been shown to have detrimental effects on population health through various pathways including health-related behaviours (19–21). These health effects may be modified by the type of welfare state regime in place and related social protection policies (22). Employment is generally associated with good health (23), while job loss or unemployment are associated with deleterious health outcomes (24), especially among men and those in their early and middle careers (23). While we observed similar findings for those unemployed prior and during the pandemic, we did not replicate the detrimental impact of job loss. However, participation in the furlough scheme was common, while participants who were no longer working during the initial stages of the pandemic were rare (∼3%) leading to low precision in estimates for this group. Nevertheless, we did not find strong evidence for the detrimental impacts on health behaviours that are normally associated with job loss, which may suggest these impacts are lessened or non-existent in the unique context of a pandemic.

While research combining results from several UK prospective studies makes a clear contribution to understanding the impact of the furlough scheme, there are limitations that should be taken into account while interpreting our findings. Firstly, we were not able to achieve full harmonisation of measures across studies. By focusing on comparable measures we also limited our scope to explore other aspects of diet, physical activity or sleep (such as frequency of snacking, specific kinds of physical activity, or sleep quality). Furthermore, outcomes were only analysed during the initial stages of the pandemic (April-July 2020) and relationships may change with subsequent changes to restrictions and growing economic uncertainty. Further research is needed to examine this as well as heterogeneity in the stable employed and furloughed groups in greater detail. The MCS cohort particularly, who were the youngest cohort studied, often had considerably different estimates from the older aged respondents in other cohorts, so there may be different mechanisms affecting this age group.

Despite being embedded in long standing studies, surveys during the pandemic were selective. We corrected most studies to being representative of their target population using weights derived for each study based on pre-pandemic information (and the GS study which did not have weights available exhibited similar estimates to the more nationally representative studies). Nevertheless, bias due to selective non-response cannot be excluded (25), especially as most studies (USOC being the exception) were weighted for non-response to COVID surveys but not for any residual non-response to the outcomes in question (among those who did participate in the overall COVID surveys). Similarly, bias due to unmeasured confounding cannot be ruled out and could be influential considering the small magnitude of the risk ratios observed. For example, there may be unobserved differences between participants whose jobs were retained, versus those who experienced furlough or job loss. Our fully adjusted models account for differences in some key pre-pandemic characteristics among employment groups. However, it is possible that our results reflect other traits of these employment groups, for example, how workers in different industries or occupational classes were responding to the pandemic, rather than being effects of furlough specifically. Adjustment for pre-pandemic characteristics may also have induced bias if there were unobserved determinants of both pre-pandemic characteristics and behaviour during the pandemic. However, we observed only minor differences between those furloughed and those who remained employed, therefore any bias due to unmeasured confounding is unlikely to change the interpretation of our findings.

Our analyses on outcomes of change in behaviour during the pandemic showed some differences from the main analyses. Specifically, they indicated that being furloughed was associated with increased fruit and vegetable consumption, hours of sleep and time spent exercising relative to maintaining stable employment. There may be several reasons for this: the change analyses included more studies, which implies a greater variability in measurement; these outcomes could have been picking up relatively minor changes in behaviour above or below the thresholds used in the main analyses and could still partially be reflecting effects of initial employment status on pre-pandemic diet, physical activity or sleep.

## Conclusions

Despite the economic disruption of the pandemic and lockdown, participants who were no longer working during the initial stages of the pandemic were rare, while much higher proportions participated in the UK CJR furlough scheme. We found that those who were furloughed exhibited broadly similar levels of health behaviours to those who remained in employment and there was some evidence of less risk for infrequent exercise. Continuation of the UK CJR furlough scheme has the potential to mitigate some of the adverse consequences of the pandemic and there was little evidence for detrimental impacts on population health behaviours. Our evidence suggests that the UK furlough scheme may be an important component of policies aiming to mitigate the determinantal effects of economic downturns and prevent exacerbation of inequalities.

## Data Availability

All datasets included in this analysis have established data sharing processes, and for most included studies the anonymised datasets with corresponding documentation can be downloaded for use by researchers from the UK Data Service. We have detailed the exact processes for each dataset in Supplementary Table S2.

## List of abbreviations

ALSPAC-G1: Avon Longitudinal Study of Parents and Children.
ALSPAC-G0: Parents of ALSPAC-G1.
BCS70: 1970 British Cohort Study.
CI: Confidence interval
CJRS: Coronavirus Job Retention Scheme.
ELSA: English Longitudinal Study of Ageing.
GS: Generation Scotland: the Scottish Family Health Study.
MCS: Millennium Cohort Study.
NCDS: 1958 National Child Development Study.
NS: Next Steps (formerly the Longitudinal Study of Young People in England).
RR: Risk Ratio.
UK: United Kingdom
USOC: Understanding Society.

## Additional Files

**Supplementary File 1: Supplementary Tables and Figures**

Format: docx

Contents : File includes supplementary information S1-S9.

**Supplementary File 2: Variable Coding**

Format: xlsx

Contents : Details of coding of variables within each study.

**Supplementary File 3: Meta-Analysis**

Format: xlsx

Contents : Full details of the meta-analytic results including study-specific estimates and weights.

## Declarations

### Ethics approval and consent to participate

We have detailed the ethical approval for each study in Supplementary Table S2.

### Consent for publication

Not applicable.

### Availability of data and materials

All datasets included in this analysis have established data sharing processes, and for most included studies the anonymised datasets with corresponding documentation can be downloaded for use by researchers from the UK Data Service. We have detailed the processes for each dataset in Supplementary Table S2.

### Competing interests

No conflicts of interest were declared by BW, JM, MJG, GDG, SP, GJG, JC, AJS, CB, RJS, DB, PP, LDH, EF, GBP. SVK is a member of the Scientific Advisory Group on Emergencies subgroup on ethnicity and COVID-19 and is co-chair of the Scottish Government’s Ethnicity Reference Group on COVID-19. NC serves on a data safety monitoring board for trials sponsored by Astra-Zeneca. ADH serves on the PHOSP COVID Cardiovascular Working group.

### Funding

This work was supported by the National Core Studies, an initiative funded by UKRI, NIHR and the Health and Safety Executive. The COVID-19 Longitudinal Health and Wellbeing National Core Study was funded by the Medical Research Council (MC_PC_20030).

Understanding Society is an initiative funded by the Economic and Social Research Council and various Government Departments, with scientific leadership by the Institute for Social and Economic Research, University of Essex, and survey delivery by NatCen Social Research and Kantar Public. The Understanding Society COVID-19 study is funded by the Economic and Social Research Council (ES/K005146/1) and the Health Foundation (2076161). The research data are distributed by the UK Data Service.

The Millennium Cohort Study, Next Steps, British Cohort Study 1970 and National Child Development Study 1958 are supported by the Centre for Longitudinal Studies, Resource Centre 2015-20 grant (ES/M001660/1) and a host of other co-funders.. The COVID-19 data collections in these four cohorts were funded by the UKRI grant Understanding the economic, social and health impacts of COVID-19 using lifetime data: evidence from 5 nationally representative UK cohorts (ES/V012789/1).

The English Longitudinal Study of Ageing was developed by a team of researchers based at University College London, NatCen Social Research, the Institute for Fiscal Studies, the University of Manchester and the University of East Anglia. The data were collected by NatCen Social Research. The funding is currently provided by the National Institute on Aging in the US, and a consortium of UK government departments coordinated by the National Institute for Health Research. Funding has also been received by the Economic and Social Research Council. The English Longitudinal Study of Ageing Covid-19 Substudy was supported by the UK Economic and Social Research Grant (ESRC) ES/V003941/1.

The UK Medical Research Council and Wellcome (Grant Ref: 217065/Z/19/Z) and the University of Bristol provide core support for ALSPAC. A comprehensive list of grants funding is available on the ALSPAC website (http://www.bristol.ac.uk/alspac/external/documents/grant-acknowledgements.pdf). We are extremely grateful to all the families who took part in this study, the midwives for their help in recruiting them, and the whole ALSPAC team, which includes interviewers, computer and laboratory technicians, clerical workers, research scientists, volunteers, managers, receptionists and nurses. The second COVID-19 data sweep was also supported by the Faculty Research Director’s discretionary fund, University of Bristol.

Generation Scotland received core support from the Chief Scientist Office of the Scottish Government Health Directorates [CZD/16/6] and the Scottish Funding Council [HR03006]. Genotyping of the GS:SFHS samples was carried out by the Genetics Core Laboratory at the Wellcome Trust Clinical Research Facility, Edinburgh, Scotland and was funded by the Medical Research Council UK and the Wellcome Trust (Wellcome Trust Strategic Award “STratifying Resilience and Depression Longitudinally” (STRADL) Reference 104036/Z/14/Z). Generation Scotland is funded by the Wellcome Trust (216767/Z/19/Z).

SVK acknowledges funding from a NRS Senior Clinical Fellowship (SCAF/15/02), the Medical Research Council (MC_UU_00022/2) and the Scottish Government Chief Scientist Office (SPHSU17). GJG acknowledges funding from the ESRC (ES/T009101/1). DB acknowledges funding from the Medical Research Council (MR/V002147/1).

### Role of funder

The funders had no role in the methodology, analysis or interpretation of the findings presented in this manuscript.

### Author Contribution Statement

Ploubidis, Katikireddi, Patalay and Chaturvedi conceptualised the study and design. Wielgoszewska. Maddock. Green, Di Gessa, Parsons, Ploubidis, Katikireddi, Griffith and Silverwood designed the methodology. Wielgoszewska. Maddock, Green, Di Gessa, Parsons, Croft, Stevenson, Booth and Griffith conducted the formal analysis. Maddock, Green, Di Gessa, Stevenson, Griffith, Chaturvedi, Howe and Fitzsimons were responsible for data curation. Wielgoszewska, Maddock. Green, Di Gessa, and Parsons wrote the original draft of the manuscript. All authors contributed to critical revision of the manuscript. Green and Di Gessa contributed to data visualisation. The project was supervised by Ploubidis and Katikireddi. Funding was acquired by Patalay, Katikireddi, Ploubidis, Silverwood, and Chaturvedi.

## Acknowledgements

The contributing studies have been made possible because of the tireless dedication, commitment and enthusiasm of the many people who have taken part. We would like to thank the participants and the numerous team members involved in the studies including interviewers, technicians, researchers, administrators, managers, health professionals and volunteers. We are additionally grateful to our funders for their financial input and support in making this research happen.

GS: Drew Altschul, Chloe Fawns-Ritchie, Archie Campbell, Robin Flaig.

ALSPAC: Daniel J Smith, Nicholas J Timpson, Kate Northstone

Understanding Society: Michaela Benzeval

MCS, NS, BCS70, NCDS: Colleagues in survey, data and cohort maintenance teams

## Notes

### Author Declarations

All details in a supplementary table

